# DMN-Targeted TMS Reduces Neural Cue-Reactivity in Individuals with Schizophrenia who Use Nicotine

**DOI:** 10.64898/2026.09.15.26363162

**Authors:** Yunong Bai, Anna Huang, Jillian G. Connolly, Sophia H. Blyth, Simon Vandekar, Baxter P. Rogers, Libby Tunison, Catie Chang, Hilary Tindle, Stephan Heckers, Roscoe O. Brady, Amy C. Janes, Heather Burrell Ward

**Author notes:** Corresponding Author Heather Burrell Ward, MD 1601 23^rd^ Ave S Nashville, TN 37212. Disclosures: The authors have no conflicts of interest to disclose Abstract: 250.

## Abstract

**Background:** Nicotine dependence is a major contributor to early mortality in individuals with schizophrenia, yet effective treatments remain limited. Drug cue-reactivity is a key contributor to nicotine use. Although transcranial magnetic stimulation (TMS) can reduce nicotine use and cue-reactivity, conventional left dorsolateral parietal (DLPFC)-targeted TMS is less effective in those with schizophrenia, highlighting the need for alternative, circuit-based targets. Because default node network (DMN) function is implicated in schizophrenia, cue-reactivity, and nicotine use, we investigated whether DMN-targeted TMS modulates cue-elicited brain activity in nicotine-using individuals with schizophrenia.

**Study Design:** 63 nicotine-using individuals (schizophrenia: n=31, non-psychosis control: n=32) participated in a randomized, crossover study comparing DMN-targeted and DLPFC-targeted TMS with pre/post-TMS neuroimaging and craving assessment. A nicotine cue-reactivity task was used concurrently with neuroimaging. Mixed-effect models were used to determine effects of TMS target and diagnosis, and model relationship between baseline craving and cue-reactivity change.

**Study Results:** At baseline, we found a Cue Reactive Network composed of regions overlapping with the DMN, including clusters in the posterior cingulate cortex, medial prefrontal cortex, and bilateral lateral occipital cortex (voxel-wise p<0.005, cluster p<0.05). Baseline cue-reactivity in schizophrenia did not differ from controls. DMN-targeted TMS decreased cue-reactivity in schizophrenia across Cue Reactive Network (estimate=-0.180, p=0.009). DLPFC-targeted TMS did not impact cue-reactivity. We did not observe an effect of either TMS target in controls. Higher baseline unprovoked craving predicted larger decrease in cue-reactivity across the Cue Reactive Network (estimate=-0.034, p=0.004).

**Conclusion:** DMN-targeted TMS is a promising intervention that modulates nicotine cue-elicited brain activity in schizophrenia.

## Introduction

Smoking is a leading contributor to early mortality in individuals with schizophrenia spectrum disorders^1^. The prevalence of nicotine dependence is 3-fold higher in individuals with schizophrenia compared to the general population^2^, and schizophrenia individuals smoke more heavily^3^. However, both pharmacological and neuromodulation treatment for nicotine dependence treatment are 50% less effective in schizophrenia^4,5^. There is a pressing need for novel treatment targets for nicotine cessation in individuals with schizophrenia.

One potential target is drug cue-reactivity, a phenomenon where drug-related cues elicit behavioral, physiological, and neural responses, such as subjective craving, sympathetic activation, and brain activity^6^. Cue-reactivity contributes to drug craving, use, and relapse^6–9^. A meta-analysis reported prospective association between cue-reactivity and drug use and relapse, with per-unit increase in cue-induced craving doubling the risk of future use^7^. In nicotine dependence, higher cue-reactivity is linked to shorter latency to smoke, increased consumption, and longer puff duration^10^. Cue-reactivity may contribute to continued smoking and relapse vulnerability^8,10^. Cue-elicited brain activity is one measurement of cue-reactivity, which correlates with unprovoked nicotine craving^9^, predicts smoking relapse^9^, and mediates the link between resting-state connectivity and cue-induced craving^11^.

Individuals with schizophrenia have demonstrated altered nicotine cue-elicited brain activity, though findings have been mixed. Potvin et al reported higher cue-induced activation in ventral medial prefrontal cortex (vmPFC) among schizophrenia smokers compared to controls^12^. Moran et al reported that compared to control smokers, smokers with schizophrenia had lower cue-induced reactivity in the bilateral superior frontal gyrus, which also negatively correlated with higher negative symptom severity^9^.

A method to alter brain activation to drug cues is through transcranial magnetic stimulation (TMS). TMS applies magnetic stimuli to a targeted brain area using either inhibitory continuous theta-burst stimulation (cTBS) or excitatory intermittent theta-burst stimulation (iTBS). It can modulate cortical excitability and ultimately induce neuroplastic changes^13^. In non-psychosis individuals with nicotine dependence, TMS targeting the dorsolateral prefrontal cortex (DLPFC) has been shown to reduce smoking behavior^14,15^, nicotine dependence^15^, cue-induced craving^15–17^ and brain reactivity to smoking cues as measured by electroencephalography (EEG)^16^. In 2020, TMS applied to the prefrontal cortex received clearance from the U.S. Food and Drug Administration for short-term smoking cessation in the general population^18^.

However, DLPFC-targeted iTBS is less effective in treating nicotine dependence in individuals with schizophrenia^19–22^, suggesting another target may be more effective in those with schizophrenia. One promising target is the default mode network (DMN), which is a functional brain network that is both active at rest and involved in goal-directed tasks requiring internally directed and/or self-referential cognition^23^. DMN consistently shows greater reactivity to smoking cues^24^ and across substance use disorders^6^, and a randomized controlled trial found that treatment-induced reduction in PCC reactivity to smoking cues was linked with decreased smoking^25^. Higher DMN connectivity correlates with higher nicotine craving in both those with and without schizophrenia^12,26^. Meanwhile, abnormal DMN connectivity and activity are also observed in schizophrenia. Individuals with schizophrenia demonstrate higher DMN connectivity at rest^27^, inadequate DMN suppression during cognitive tasks^28^, and brain-wide dysconnectivity involving the DMN characterized by increased DMN-salience network connectivity and mixed changes in DMN-executive control network^29^. Acute nicotine administration normalized DMN hyperconnectivity only in those with schizophrenia^30^. Higher connectivity from lateral parietal node of DMN to the rest of DMN was associated with higher likelihood of lifetime smoking only in psychosis individuals, with a diagnosis-by-connectivity interaction^31^. Collectively, these findings indicate that while DMN function is broadly implicated in nicotine dependence, distinct DMN connectivity patterns are associated with nicotine use in individuals with schizophrenia, suggesting that targeting DMN may be especially effective in schizophrenia populations. The lateral parietal cortex is an ideal target for DMN neuromodulation given its superficial location and reliable identification across individuals^32^.

We aimed to test if modulating DMN via TMS would affect neural cue-reactivity. To determine the specificity of DMN modulation, we compared the impact of two different TMS interventions (DMN-targeted cTBS and DLPFC-targeted iTBS) on cue-elicited brain activity in two different groups of people who use nicotine (schizophrenia and non-psychosis control). Using measurements of blood-oxygen-level-dependent (BOLD) signal while viewing Nicotine or Neutral images before and after each TMS intervention, we tested 1) whether cue-reactivity in schizophrenia differed from non-psychosis control at baseline, and 2) whether cue-reactivity in schizophrenia responded differently to different TMS interventions compared to non-psychosis control. As an exploratory analysis, we also tested if TMS-induced cue-reactivity change can be predicted by baseline craving.

## Methods

### Participants

Sixty-three participants provided neuroimaging data for analysis: 1) Schizophrenia (n=31) and 2) Non-psychosis control (n=32) (Table 1, Figure 1, Supplemental Figure 1, NCT06389266). Individuals in the schizophrenia group had a DSM-V diagnosis of schizophrenia or schizoaffective disorder. Individuals in the non-psychosis control group had no lifetime history of psychosis but could have other DSM-V diagnoses, such as depression and anxiety (Table 1). Participants provided written informed consent. Details in supplement.

**Figure 1.**
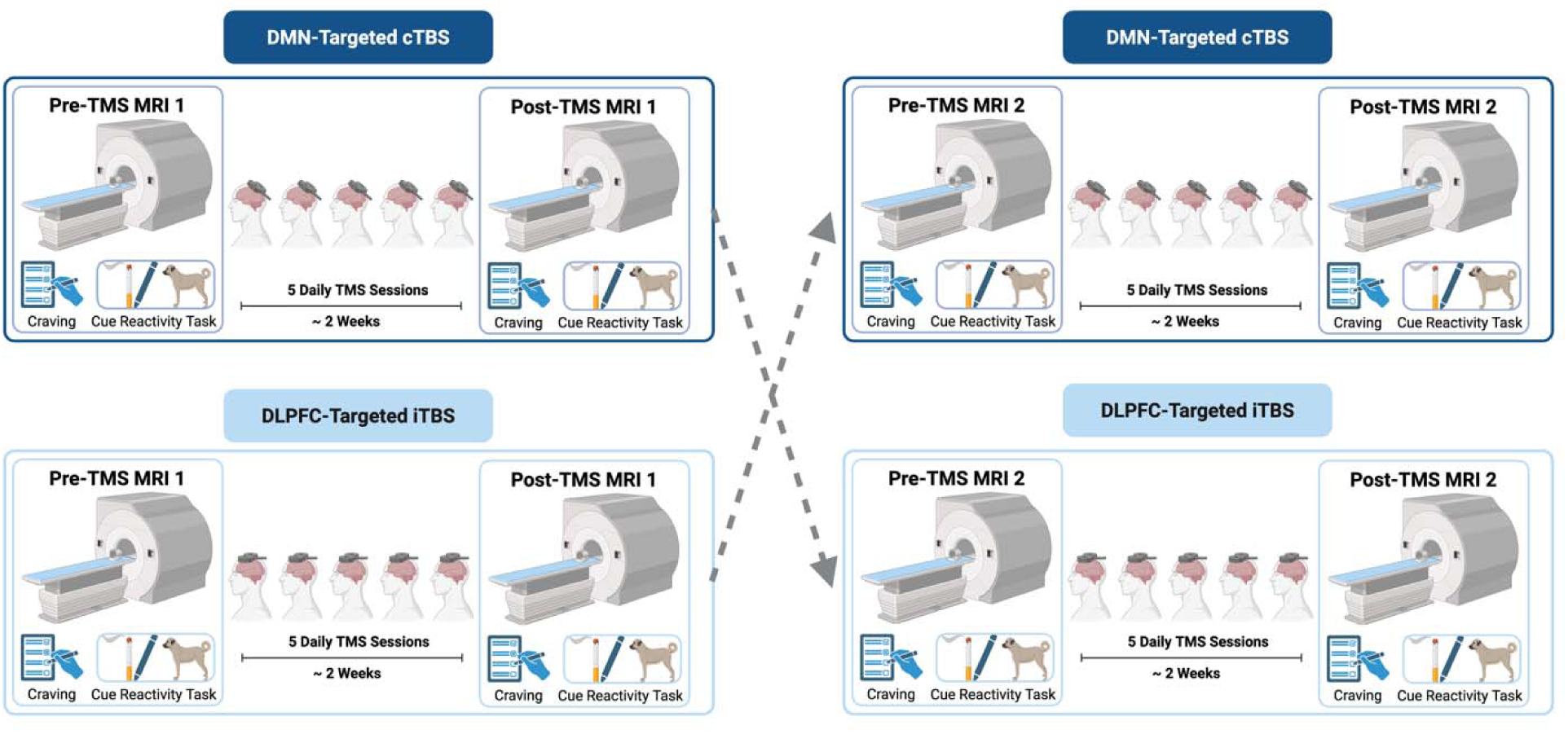
Study Design. 63 individuals who use nicotine participated in this randomized, crossover transcranial magnetic stimulation (TMS) study comparing DMN (default mode network)-targeted TMS to left dorsolateral prefrontal cortex (DLPFC)-targeted TMS. Participants were enrolled in two groups: schizophrenia (n=30) and non-psychosis control (n=32). Participants received 5 daily sessions of 1) DMN-Targeted cTBS (continuous theta burst stimulation) and 2) L DLPFC-Targeted iTBS (intermittent theta burst stimulation) in a randomized, crossover design. We used functional magnetic resonance imaging (fMRI) to measure nicotine cue-elicited brain activity before and after completing each TMS protocol. Participants were shown nicotine (cigarette or vape), neutral, and target cues while during fMRI scan. Individuals who smoke cigarette were shown cigarette cues, and those who vape were shown vaping cues. Craving was measured using the Visual Analog Scale immediately before cues were presented. Created with BioRender.com.

**Table 1.** Demographics.

|  |  | Schizophrenia<br>(n=31) | Control<br>(n=32) | P-value | Test |
| --- | --- | --- | --- | --- | --- |
| Demographics |  |  |  |  |  |
|  | Age (years), mean (SD) | 32.81 (11.43) | 37.13 (13.11) | 0.168 | T-test |
|  | Gender (female), n (%) | 16.13 | 34.37 | 0.169 | Chi-square |
|  | Race, n (%) |  |  | 0.005* | Chi-square |
|  | Black or African American | 12 (38.7) | 1 (3.8) |  |  |
|  | White | 16 (51.6) | 28 (87.5) |  |  |
|  | Multi-Racial | 2 (6.5) | 2 (6.2) |  |  |
|  | Other | 1 (3.2) | 1 (3.8) |  |  |
|  | BPRS Total, mean (SD) | 40.5 (12.2) | 29.6 (5.3) | <0.001* | T-test |
|  | CPZ Equivalents, mean (SD) | 207.26 (345) | 14.06 (79.5) | 0.004* | T-test |
| Nicotine Use History |  |  |  |  |  |
|  | Type, n (%) |  |  | 0.901 | Chi-square |
|  | Combustible | 16 | 15 |  |  |
|  | Noncombustible | 15 | 17 |  |  |
|  | FTND, mean (SD) | 4.43 (1.96) | 2.06 (2.10) | 0.001* | T-test |
|  | PSECDI, mean (SD) | 10.43 (4.91) | 9.91 (5.01) | 0.798 | T-test |
| Non-Psychosis Psychiatric History, n |  |  |  |  |  |
|  | Depression | - | 14 |  |  |
|  | Anxiety | - | 15 |  |  |
|  | ADHD | - | 7 |  |  |
|  | Bipolar Disorder | - | 2 |  |  |
|  | PTSD | - | 2 |  |  |
|  | Borderline Personality Disorder | - | 1 |  |  |
|  | Adjustment Disorder | - | 1 |  |  |

ADHD: attention deficit and hyperactivity disorder; BPRS: Brief Psychiatric Rating Scale; FTND: Fagerstrom Test for Nicotine Dependence; PSECDI: Penn State Electronic Cigarette Dependence Index; PTSD: post-traumatic stress disorder.

### Nicotine Assessments

Nicotine dependence was assessed using the Fagerstrom Test for Nicotine Dependence (FTND)^33^ and Penn State Electronic Cigarette Dependence Index (PSECDI)^34^.

### MRI Acquisition

Imaging data were collected on 3.0-T Philips MR7700 or Achevia MRI scanner (Philips Healthcare, Andover, MA). Participants completed MRI scans the week before and after TMS intervention. As the durability of 5 TMS sessions is unknown, post-TMS scans were scheduled within 8 days after TMS (mean 4.39 (SD 1.61) days). 1-mm^3^ T1-weighted anatomical scans and 3 blocks of cue-reactivity task scans were acquired (TR 2000ms, TE 28.0ms, flip angle 90 degrees, field of view = 240mm, 38 slices, 3-mm^3^ voxels, anterior to posterior phase-encoded).

### Nicotine Cue-reactivity Task

The in-scanner cue-reactivity task was similar to previously published work^11,35,36^. While in the MRI, participants were shown 30 nicotine (cigarette or vape, depending on use history), 30 neutral, and 6 target images divided evenly across 3 blocks lasting 5min 18s each. Images were presented for 4s in a pseudorandom order (with no more than two of the same picture type occurring in a row). Participants viewed smoking/vaping and matched neutral images and pressed a button when an animal target image was displayed. Participants rated craving using 0 to 10 Visual Analog Scale before cue presentation. The task was administered at each of the 4 MRIs without repeating images. Details in supplement.

### MRI Processing

Task scans were preprocessed in SPM12 (Wellcome Department of Cognitive Neurology, London, UK) in Matlab 23.2.0 (R2023b) Update 10 (MathWorks, Inc., Natick, MA). All task scans went through quality control procedures (see Supplement). After quality control, data from 21 schizophrenia and 27 non-psychosis control participants were included in analysis (Supplemental Figure 1). Individual subject data were analyzed using general linear models with regressors for Nicotine, Neutral, and Target stimuli, 6 motion parameters. Each stimulus was modeled at the onset of each image and had a duration of 4 seconds. In group analysis, we performed one-sample t-tests on first-level Nicotine>Neutral contrast maps from baseline task scans only (collected prior to receiving any TMS intervention) across all participants to identify baseline Nicotine cue-reactive regions. Voxel-level threshold was p<0.005. Cluster-level threshold was FDRp<0.05. Regions from the thresholded second-level Nicotine>Neutral reactivity map were defined as the Cue Reactive Network. SPM MarsBar toolbox was used to segment individual clusters from the Cue Reactive Network. To explore whether the effects of TMS were driven by a specific cluster, we conducted a follow-up analysis in which each region of interest (ROI) was examined separately. Each cluster was defined as a region-of-interest (ROI), including the posterior cingulate cortex (PCC), medial prefrontal cortex (mPFC), left lateral occipital, right lateral occipital. Using MarsBar, we extracted ROI-wise beta values from first-level outputs (Nicotine-Baseline, Neutral-Baseline, Nicotine>Neutral) across the Cue Reactive Network and for each ROI on every task scan. Details in supplement.

### TMS Protocol

In this randomized, crossover trial, individuals received five daily sessions of 1) DMN-Targeted cTBS and 2) left DLPFC-Targeted iTBS with pre-/post-TMS neuroimaging. TMS order was randomized, and interventions were separated by a two-week washout period. DMN-Targeted cTBS (600 pulses, 100% AMT) was applied to an individualized left parietal DMN target (see *Individualized DMN Target* in Supplement). Left DLPFC-Targeted iTBS (600 pulses, 100% AMT) was anatomically targeted using MNI coordinates for the average 5cm rule (x = -41, y = +16, z = +54)^37^. Details in supplement.

### Statistical Analysis

We performed a staged analysis to first investigate group differences and TMS effect on cue-reactivity across the whole Cue Reactive Network, then investigated group differences and TMS effect within each ROI in this network. All statistical analyses were performed in RStudio version 4.5.2. P-values were adjusted using FDR across all ROIs using the Benjamini-Hochberg procedure. Results with α<0.05 were designated significant.

### Pre-Treatment Group Differences

To investigate group differences in cue-reactivity prior to TMS, we used random-intercept linear mixed-effect (LME) to model reactivity contrast (Nicotine>Baseline, Neutral>Baseline) using diagnosis-by-cue-type interaction, controlling for age and random intercepts for subject, using data collected prior to the first TMS session. Only baseline scans prior to any TMS treatment were included. Diagnostic differences in marginal means were evaluated for each cue type using estimates, confidence intervals, and t-tests. Two sensitivity analyses were performed to investigate effects of medication and symptom severity by fitting two LME models within schizophrenia to model cue-reactivity using (1) chlorpromazine equivalent dose and (2) Brief Psychiatric Rating Scale (BPRS), controlling for age and random intercepts for subject.

### TMS Effect

To study the effect of TMS on cue-reactivity, we used LME to model TMS-induced change in Nicotine>Neutral reactivity (post – pre) using diagnosis-by-TMS-type interaction, controlling for age, pre-TMS Nicotine>Neutral reactivity, and random intercepts for subject. Estimated marginal means of TMS-induced reactivity change was calculated for each group and TMS target. Pre-TMS measurement is specific to each block of TMS treatment. Only scans with both pre-TMS and post-TMS data available were included in this analysis. Marginal means for each diagnosis and TMS type combination were calculated to evaluate if TMS-induced change was different from 0. We performed two sensitivity analyses: (1) adding TMS order (DMN first or L DLPFC first) as another covariate, and (2) whether Nicotine>Neutral reactivity changed between the start of each TMS block (pre-TMS2 – pre-TMS1, “baseline stability”), and between the end of the first TMS block and the start of the second TMS block (pre-TMS – post-TMS1), “washout”).

### Predicting TMS-Induced Cue-reactivity Change

We investigated whether pre-TMS behavioral measurements would predict TMS-induced change in Nicotine>Neutral reactivity. LME was used to model TMS-induced change in reactivity using pre-TMS in-scanner unprovoked self-rated craving, controlling for age, pre-TMS reactivity, and random intercepts for subject. This analysis was done collapsing all groups and treatment targets.

## Results

The voxel-wise, group-level analysis identified a Cue Reactive Network (Figure 2A). Regions in this network were more reactive to Nicotine cues compared to Neutral cues at baseline (voxel-level p<0.005, cluster p<0.05). Four significant clusters were identified within the Cue Reactive Network: Posterior Cingulate (PCC, peak voxel at MNI [0,-36,32]; Figure 2B), Medial Prefrontal Cortex (mPFC, peak voxel at MNI [0,38,4]; Figure 2C), Left Lateral Occipital (peak voxel at MNI [-40,-54,-14]; Figure 2D), Right Lateral Occipital (peak voxel at [50,-62,2]; Figure 2E).

**Figure 2.**
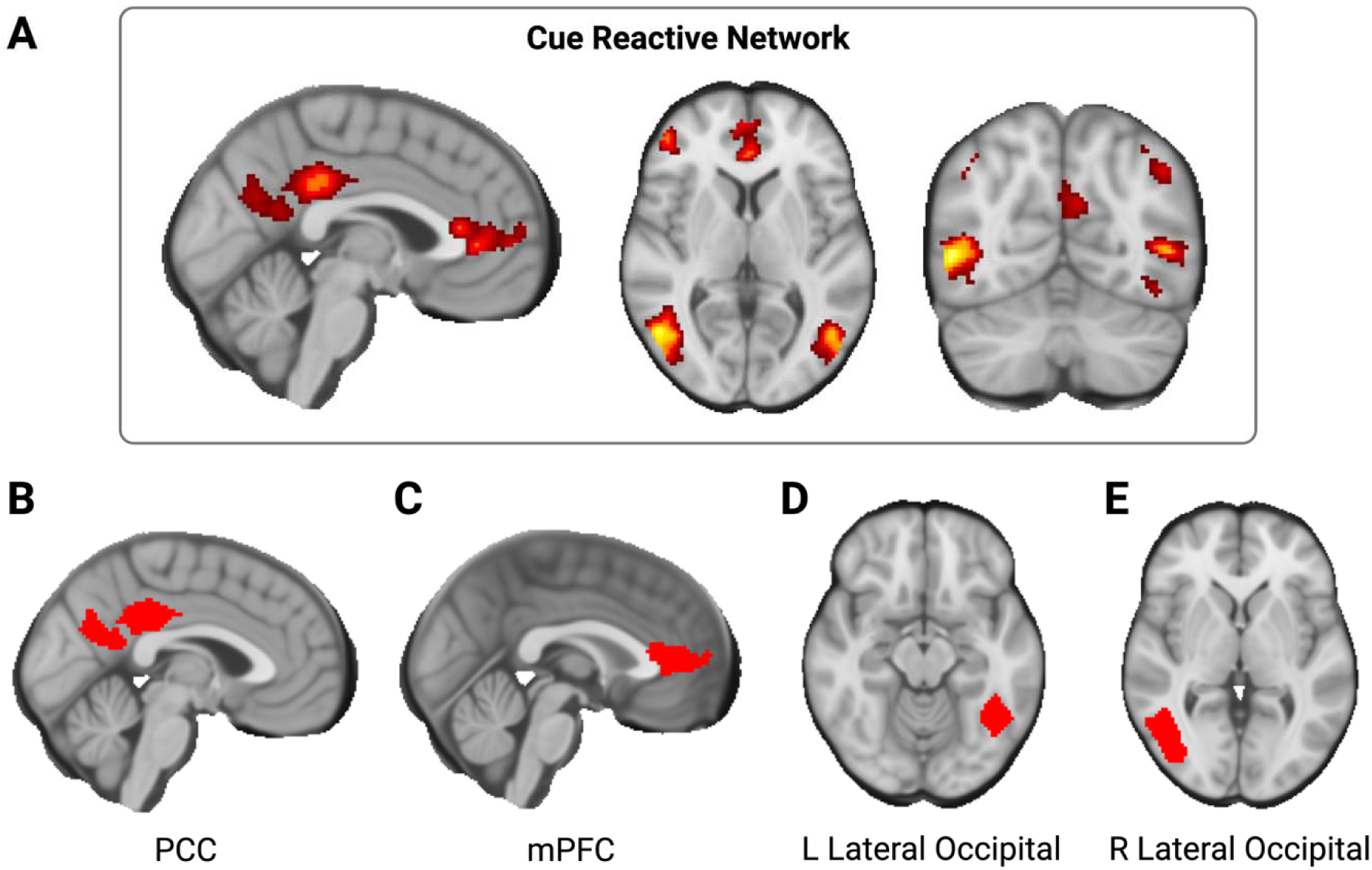
Voxel-Wise, Group-Level Cue-Reactivity Analysis Identified the Cue Reactive Network. Regions in this network were more reactive to Nicotine cues compared to Neutral cues at baseline (voxel-level p<0.005, cluster p<0.05). Four significant clusters were identified within the Cue Reactive Network: posterior cingulate (PCC, peak voxel at MNI [0,-36,32]), medial prefrontal cortex (mPFC, peak voxel at MNI [0,38,4]), left lateral occipital (peak voxel at MNI [-40,-54,-14]), right lateral occipital (peak voxel at [50,-62,2]). Created with BioRender.com.

### Pre-Treatment Cue-reactivity Did not Differ by Diagnosis or Correlate with Symptom Severity

We modeled cue-reactivity using diagnosis-by-cue-type interaction, controlling for age. Analyses were performed on the entire Cue Reactive Network and ROIs identified from the previous step. There was no significant interaction, and reactivity to Nicotine or Neutral cues did not differ between schizophrenia and non-psychosis control in any region (p>0.05; Supp. Table 1). Pre-treatment Nicotine>Neutral contrast did not correlate with age, chlorpromazine equivalents, BPRS total, or BRPS negative (p>0.05); see Supp. Table 2).

### DMN-Targeted cTBS Decreased Cue-Reactivity in Schizophrenia

We modeled TMS-induced reactivity change using diagnosis-by-TMS-type interaction, controlling for age. Diagnosis-by-TMS-type interaction showed trend-level relationship with change in Cue Reactive Network reactivity (estimate=-0.196, SE=0.117, Z=-1.678, df=56, p=0.099). Estimated marginal means of each group and TMS target showed that in schizophrenia, DMN-targeted cTBS significantly decreased Nicotine>Neutral contrast in the entire Cue Reactive Network (estimate=-0.180, SE=0.067, Z=-2.698, DF=56, p=0.009; Figure 3). This decrease was driven by decreased reactivity to Nicotine cues (Nicotine>Baseline: estimate=-0.135, SE=0.062, Z=-2.157, DF=56, p=0.035; Figure 3), while reactivity to Neutral cues did not change (Neutral>Baseline: estimate=0.058, SE=0.059, Z=0.981, DF=55.6, p=0.33; Figure 3). A follow up analysis indicated this was driven by changes in the PCC (estimate=-0.246, SE=0.095, Z=-2.589, DF=56, p=0.012, FDRp=0.036; Supp. Figure 2B, Supp. Table 3), and Left Lateral Occipital (estimate=-0.161, SE=0.066, Z=-2.433, DF=56, p=0.018, FDRp=0.036; Supp. Figure 2C, Supp. Table 3); mPFC cue-reactivity also showed trend-level changes (estimate=-0.145, SE=0.080, Z=-1.803, DF=56, p=0.077, FDRp=0.077; Supp. Table 3). In non-psychosis control, DMN-targeted cTBS did not change Nicotine>Neutral contrast in Cue Reactive Network (p>0.05). DLPFC-targeted iTBS did not change Nicotine>Neutral contrast in Cue Reactive Network (p>0.05) in either group. Sensitivity analyses showed no effect of TMS order (p>0.05, Supp. Table 4), cue-reactivity did not change during TMS washout (pre-TMS2 – post-TMS1; p>0.05, Supp. Table 5) and remained stable at TMS baseline (pre-TMS2 – pre-TMS1; p>0.05, Supp. Table 5).

**Figure 3.**
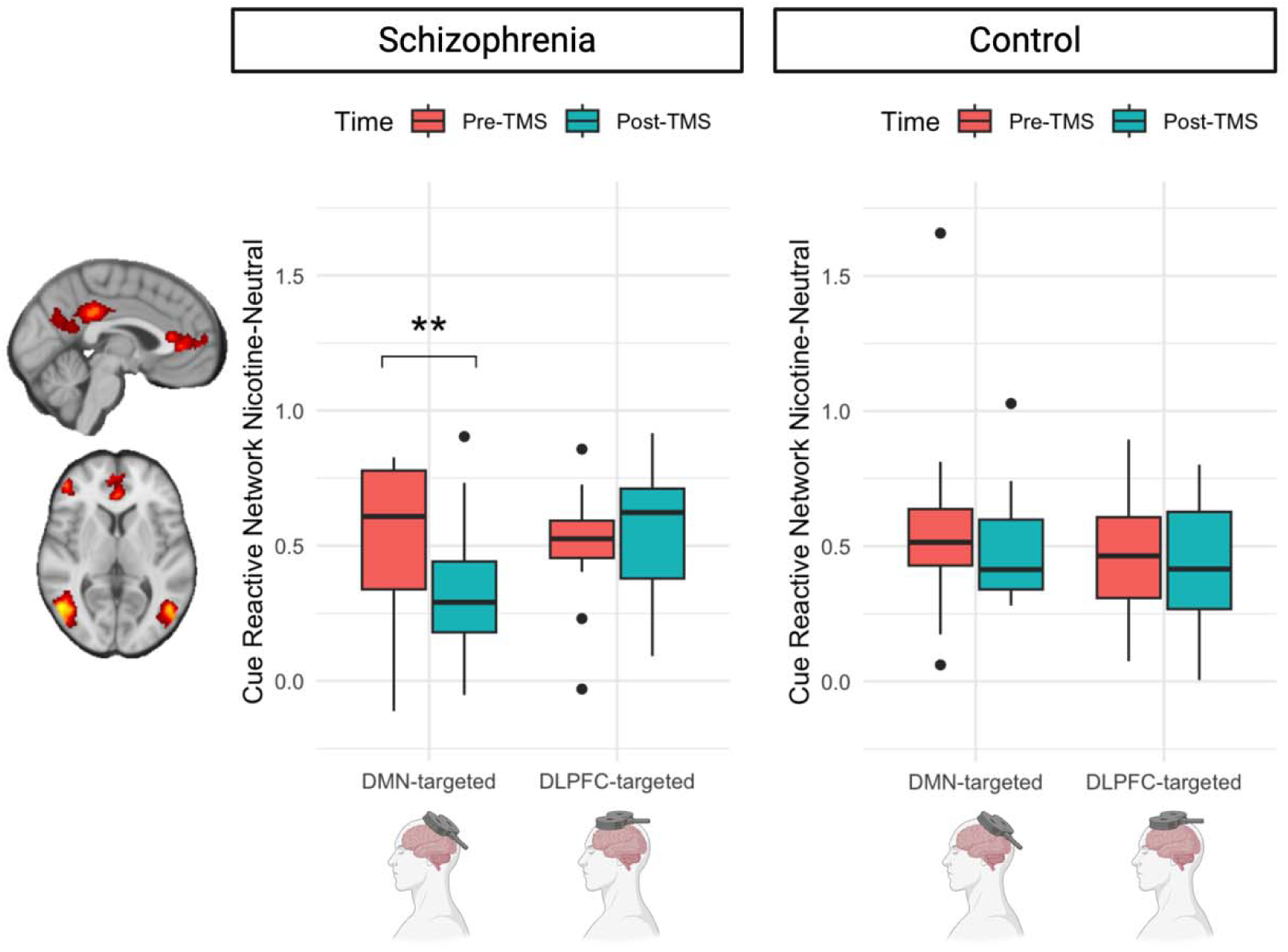
DMN-Targeted cTBS Decreased Cue Reactivity in Schizophrenia Across the Cue Reactive Network. We modeled TMS-induced cue-reactivity change using diagnosis-by-TMS-type interaction, controlling for age. Estimated marginal means indicated that only the schizophrenia/DMN-targeted cTBS group exhibited significant changes in cue-reactivity. DMN-targeted cTBS significantly decreased Nicotine>Neutral contrast in Cue Reactive Network in schizophrenia (estimate=-0.180, p=0.009), while no effect was observed in the control group. DLPFC-targeted iTBS did not change Nicotine>Neutral contrast in Cue Reactive Network in either group. Created with BioRender.com.

### Higher Pre-TMS Subjective Craving was Associated with Larger TMS-Induced Decreases in Nicotine>Neutral Reactivity

We used pre-TMS subjective craving prior to cue exposure to predict TMS-induced change (post-TMS Nicotine>Neutral – pre-TMS Nicotine>Neutral) in the Cue Reactive Network, controlling for pre-TMS Nicotine>Neutral contrast and age. Greater pre-treatment subjective craving was associated with lower post-TMS brain reactivity to nicotine vs neutral cues within the entire Cue Reactive Network (estimate=-0.034, SE=0.011, Z=-3.021, DF=55, p=0.004; Figure 4A, Supp. Table 6) and each of the Cue Reactive Network subregions (FDRp’s<0.05, Figure 4A-E, Supplemental Table 6).

**Figure 4.**
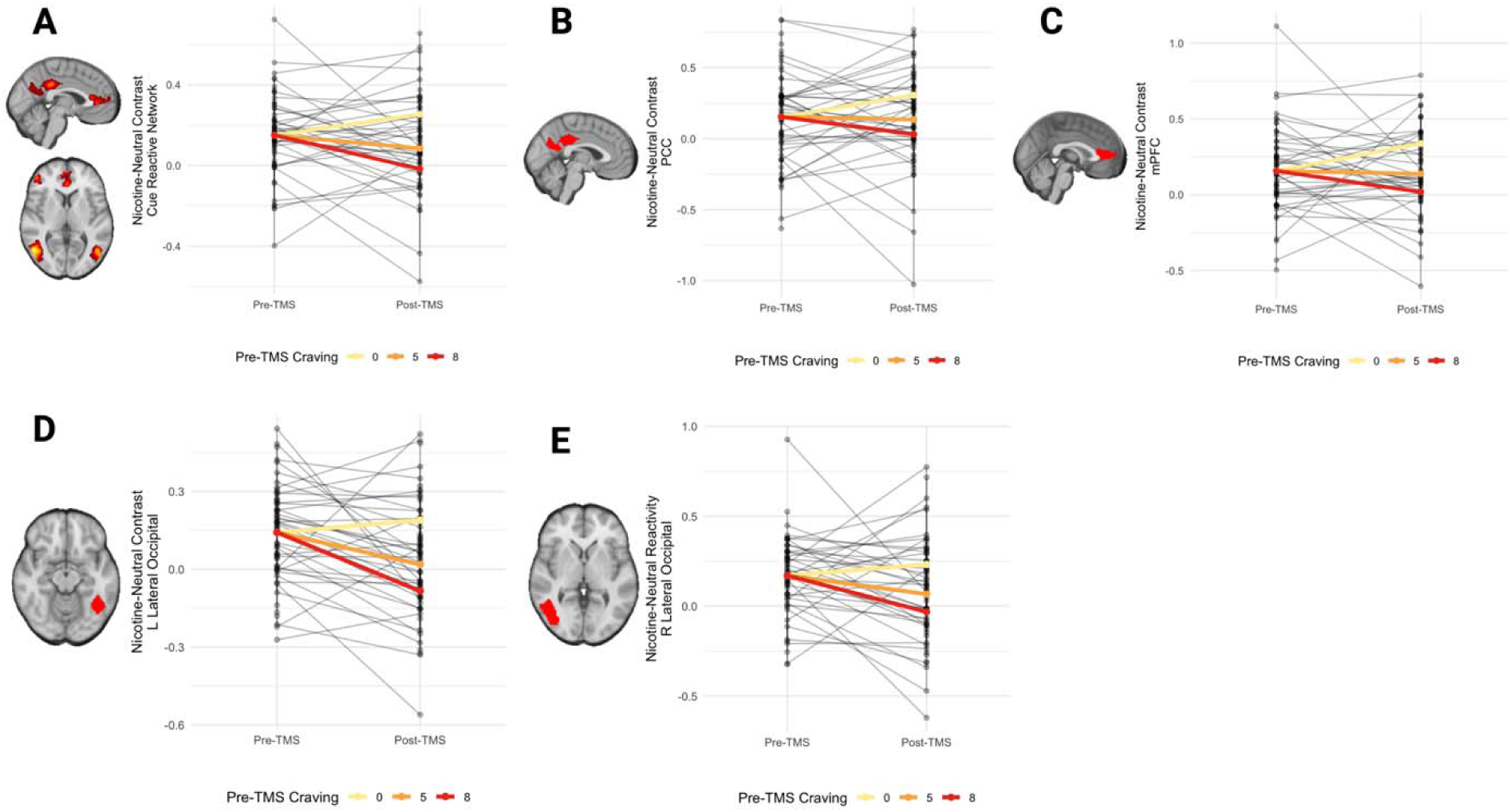
Change in Cue-Reactivity was Predicted by Pre-TMS Unprovoked Craving. Pre-TMS craving negatively associated with post-TMS Nicotine>Neutral contrast in Cue Reactive Network (estimate=-0.034, p=0.004; Figure 4A), posterior cingulate (PCC, estimate=-0.034, p=0.039, FDRp=0.039; Figure 4B), medial prefrontal cortex (mPFC, estimate=-0.040, p=0.003, FDRp=0.007; Figure 4C), left lateral occipital (estimate=-0.034, p=0.002, FDRp=0.007; Figure 4D), right lateral occipital (estimate=-0.033, p=0.029, FDRp=0.039; Figure 4E). To interpret this relationship, we estimated simple slopes (marginal trends) of time predicting Nicotine>Neutral contrast change from our mixed effects model. Slopes were computed separately by group at the 10^th^, 50^th^, and 80^th^ percentiles of pre-TMS craving (0, 5, 8). Higher pre-TMS craving predicted larger decrease in cue-reactivity across Cue Reactive Network given average pre-TMS cue-reactivity and age. Created with BioRender.com.

## Discussion

Using data from a randomized, crossover trial evaluating two TMS interventions in schizophrenia and non-psychosis control participants, we compared baseline differences and treatment effects on cue-reactivity between the groups. Prior to TMS, we identified a Cue Reactive Network composed of regions more reactive to nicotine versus neutral cues, which did not differ by diagnosis or correlated with psychosis symptom severity. DMN-targeted cTBS decreased cue-reactivity only in those with schizophrenia, while we did not find evidence that DLPFC-targeted iTBS changed cue-reactivity in either group. Across both groups and treatment targets, higher pre-TMS unprovoked craving was associated with a greater reduction in brain reactivity to nicotine cues following TMS.

This is the first study to compare the effects of two TMS interventions on cue-elicited brain activity in two different diagnostic groups. Consistent with our hypotheses, DMN-targeted cTBS reduced brain activation to nicotine cues in schizophrenia, and we did not observe this effect in the control group. While we observed a brain-based effect of DMN-targeted cTBS in schizophrenia with only five sessions, it is likely that more TMS sessions would produce a stronger effect. Future studies should up-scale DMN-targeted cTBS beyond five sessions. These promising findings suggest that further research investigating DMN-targeted cTBS in schizophrenia is warranted to determine if it could be effective to reduce actual nicotine use and promote nicotine cessation.

We found that the Cue Reactive Network is located in midline DMN and bilateral lateral occipital cortices, and that TMS-induced changes in Nicotine>Neutral reactivity across Cue Reactive Network in schizophrenia was driven by changes in PCC and Left Lateral Occipital. This is consistent with canonical drug cue-reactivity regions reported by several meta-analyses^6,38,39^. Specifically, PCC and mPFC reactivity were a stable, transdiagnostic endophenotype of addiction across cue reactivity studies^39,40^. The midline DMN has extensive functional connectivity across the brain^41^ and integrates cortical information to facilitate self-related decision making^42,43^. Others have shown that a nicotine-related cue-induced increase in anterior DMN activity correlates with the expectation of immediate smoking^44^ and nicotine dependence^45^, while Drug>Food activity in posterior DMN was theorized to suggest greater internally-directed attention triggered by drug cues^46^. Our findings indicate that activity of canonical drug cue-reactivity regions can be modulated by DMN-targeted cTBS in schizophrenia nicotine users and further demonstrate its target engagement.

Additionally, we found that in both groups and TMS targets, higher pre-treatment subjective craving prior to cue exposure predicted larger TMS-induced decrease in cue-reactivity across all Cue Reactive Network clusters. This finding introduces nuances to prior studies that reported higher pre-treatment both unprovoked and cue-induced craving predicted greater relapse risk following nicotine replacement therapy^47,48^. Cue-reactivity has been consistently associated with clinical outcomes^6–10,49,50^. These studies support that treatment can modify cue-reactivity and generate clinical improvement. While individuals with higher pre-treatment craving typically have worse prognosis, our finding suggests that TMS is a promising nicotine cessation treatment especially in patients who may respond poorly to existing treatment options.

Our study has several major strengths. The crossover design allowed us to compare responses to two interventions within each participant. Using an active control (DLPFC-targeted iTBS), we could direct compare a novel intervention against an established one. We defined personalized standard anatomical targets for TMS and located the target using MRI-guided neuronavigation, which is the gold-standard for implementing personalized TMS protocols^51^. Furthermore, participants smoked and/or vaped, allowing for investigating the impact of both combustible and noncombustible nicotine use. Others have shown that smoking and vaping stimuli did not evoke different responses in brain reactivity or cue-induced subjective craving^11^. Including participants who vape nicotine is critical, as the prevalence of vaping is increasing,^52^ making nicotine cessation treatment development for this cohort clinically relevant. Additionally, participants in the control group were allowed to have non-psychosis psychiatric diagnoses, which is more representative of nicotine users in the general population. Notably, the activation patterns found in the current populations are consistent with prior literature^6^, suggesting that cue reactivity is not profoundly impacted by including other psychiatric diagnoses in a naturalistic manner.

There are several limitations. There is no sham control, so we cannot compare TMS effects against treatment-as-usual. Compared to the active control, DMN-targeted cTBS demonstrated differential effects with target engagement, making the effect less likely to be contributed to trial participation. Additionally, our sample did not exhibit baseline cue-reactivity differences by diagnosis. Our relatively small sample size may be underpowered to detect baseline differences. The lack of differences could also be attributed to the inclusion of non-psychosis psychiatric diagnoses in the control group.

In conclusion, DMN-targeted cTBS is a novel intervention for nicotine dependence that modulates nicotine cue-elicited brain activity in schizophrenia. These findings represent a paradigm-shift in therapeutic development by establishing consistent mechanistic evidence for DMN involvement in nicotine dependence in those with schizophrenia, providing a strong rationale for the development DMN-targeted cTBS as a nicotine cessation treatment for schizophrenia.

## Data Availability

All data produced in the present study are available upon reasonable request to the authors

## Acknowledgement

The authors would like to thank the participants in this study.

