## Supplement for "DMN-Targeted TMS Reduces Neural Cue-Reactivity in Individuals with Schizophrenia who Use Nicotine"

**Supplemental Methods**

*Participants:* Ninety individuals aged 18-65 who use nicotine were enrolled. Participants were recruited in two groups: 1) Schizophrenia and 2) Non-psychosis control (NCT06389266). 63 participants completed the protocol and provided complete data for analysis. Individuals in the schizophrenia group had a diagnosis of schizophrenia or schizoaffective disorder (n=30) confirmed by DSM-V SCID interview^1^ and clinical information obtained from outpatient psychiatric providers. Individuals in the non-psychosis control group had no lifetime history of psychosis (n=32) confirmed by DSM-V SCID Part B interview^1^. Non-psychosis psychiatric diagnoses for the NC group were obtained from their medical records. Information regarding non-psychosis psychiatric diagnoses for schizophrenia individuals was not collected. For one month prior to enrollment, individuals received outpatient care, with no hospitalizations or changes to their psychiatric medication regimens. Individuals were excluded if they had DSM-V intellectual disability, substance use disorder (other than nicotine) in the past 3 months confirmed by DSM-V SCID Part E interview, a progressive or genetic neurologic disorder, history of significant head trauma, history of seizures or neurosurgical procedures, implanted devices, gross organic pathology on neuroimaging, contraindications to MRI or TMS, or current pregnancy. All participants provided written informed consent in accordance with the Vanderbilt University Medical Center Institutional Review Board.

*Nicotine Cue-reactivity Task:* In the MRI, participants were shown 30 nicotine (cigarette or vape), 30 neutral, and 6 target images divided evenly across 3 blocks lasting 5min 18s each. Different types of stimuli were interleaved in a given block. Images were presented for 4s in a pseudorandom order (with no more than two of the same picture type occurring in a row). Participants who smoked completed a version of the task with smoking-related images. Participants who vaped completed an analogous version with vaping-related images. Participants viewed smoking/vaping and matched neutral images (e.g., hands holding cigarettes/vapes or neutral objects; faces in a smoking/vaping or neutral context) and pressed a button when an animal target image was displayed. Participants rated craving using 0 to 10 Visual Analog Scale before cue presentation. The task was administered at each of the 4 MRI sessions without repeating images. To standardize nicotine use and ensure craving, participants had at least 60 minutes between last nicotine use and cue-reactivity assessment.

*MRI Processing*: Anatomical images were segmented into grey matter, white matter, and cerebral spinal fluid with the Computational Anatomy Toolbox 12 (CAT12, version 12.5; [http://www.neuro.uni-jena.de/cat/](https://nam12.safelinks.protection.outlook.com/?url=http%3A%2F%2Fwww.neuro.uni-jena.de%2Fcat%2F&data=05%7C01%7Cheather.b.ward%40vumc.org%7C53146c81d5534edf7b5208db2189eebc%7Cef57503014244ed8b83c12c533d879ab%7C0%7C0%7C638140648518151829%7CUnknown%7CTWFpbGZsb3d8eyJWIjoiMC4wLjAwMDAiLCJQIjoiV2luMzIiLCJBTiI6Ik1haWwiLCJXVCI6Mn0%3D%7C3000%7C%7C%7C&sdata=Fsce8FRawoYi0jL0FjzH%2BUsdG8P5cAj4Wzyuk79pHuo%3D&reserved=0)). Task fMRI scans were preprocessed in SPM12 (Wellcome Department of Cognitive Neurology, London, UK) in Matlab 23.2.0 (R2023b) Update 10 (MathWorks, Inc., Natick, MA). Task scans were (1) realigned to a mean scan, (2) coregistered through the mean scan to a skull-stripped anatomical image, (3) normalized to MNI152 template space using nonlinear warping regularization, and (4) smoothed using a 6mm FWHM Gaussian kernel. All task scans went through a quality assurance procedure. Scans were excluded if participants moved more than 1 voxel (3 mm), had severe imaging artifacts on visual inspection, or substantial truncation at the vertex due to incomplete image acquisition. In first-level analysis, fixed-effect general linear models were fitted on task scans within each participant’s MRI session to estimate voxel-wise beta for contrasts (Nicotine>Baseline, Neutral>Baseline, Nicotine>Neutral). Six motion parameters (6 dimensions: x, y, z, yaw, pitch, roll) were included as covariates in the model. In second-level analysis, we performed one-sample t-test on first-level Nicotine>Neutral contrast maps from baseline task scans only (collected prior to receiving any TMS intervention) to identify Nicotine cue-reactive regions at baseline. Implicit mask was applied to restrict model estimation within voxels present on all scans. Explicit mask was created by selecting voxels with image signals from at least 90% of scans. Explicit mask was applied to limit cluster comparison within imaged regions. Voxel-level threshold was p<0.005. Cluster-level correction for multiple comparisons was done using FDR. Cluster-level threshold was FDRp<0.05. Regions from the thresholded second-level Nicotine>Neutral reactivity map was defined as the Cue Reactive Network, which overlapped with parts of the DMN and the visual system. We used the SPM MarsBar toolbox to segment individual clusters from the Cue Reactive Network. Each cluster was defined as a region-of-interest (ROI). Five ROIs were produced, including the posterior cingulate cortex (PCC), medial prefrontal cortex (mPFC), left lateral occipital (LLO), right lateral occipital (RLO). Using MarsBar, we extract ROI-wise beta from first-level outputs (Nicotine>Baseline, Neutral>Baseline, Nicotine>Neutral) across the Cue Reactive Network and for each ROI on every task scan.

*TMS Protocol:* In this randomized, crossover trial, individuals received 5 daily sessions of 1) DMN-Targeted cTBS and 2) L DLPFC-Targeted iTBS with pre-/post-TMS neuroimaging. TMS order was randomized, and interventions were separated by a two-week washout period. DMN-Targeted cTBS (600 pulses, 100% AMT) was applied to an individualized left parietal DMN target (see *Individualized DMN Target* below). L DLPFC-Targeted iTBS (600 pulses, 100% AMT) was anatomically targeted using MNI coordinates for the average 5cm rule for scalp-based DLPFC targeting (x = -41, y = +16, z = +54)^2^ with Brainsight neuronavigation software (Rogue Research, Inc.). TMS was applied using a MagPro X100 stimulator and an active figure-of-8 coil (Cool B65, MagVenture, Denmark) held tangentially to the scalp with the handle at 45 degrees. TMS was applied in the standard theta-burst pattern (3 pulses at 50-Hz repeated at a rate of 5-Hz)^3^. Details in supplement.

*TMS Protocol Motor Threshold Determination:* Participants had motor threshold determination at their first TMS visit. Single pulse and repetitive stimulation was performed with a MagPro stimulator (MagVenture, Denmark) equipped with a biphasic figure-of-eight coil. To obtain an active motor threshold, single pulses were used in the following manner: Electromyographic activity (EMG) was recorded using surface electrodes attached to the skin to measure motor evoked potentials (MEP) during the motor threshold assessment. The TMS coil was placed on the scalp. Single TMS pulses were applied over the hand area of the left motor cortex and individually localized for each participant based on the optimal position for eliciting a motor evoked potential. Neuronavigation (Brainsight, Rogue Research, Inc.) was used to record the motor ‘hot-spot.’ Resting and active motor threshold (RMT; AMT) were obtained by following recommendations from the International Federation of Clinical Neurophysiology.

*Individualized DMN Target*: To modulate the DMN, we selected a personalized target in the left lateral parietal region of the DMN. This target was selected based on prior work in healthy populations showing that applying TMS to this DMN node effectively modulated DMN connectivity^4^. To identify an individualized DMN map for TMS targeting, a standard DMN template^5^ was warped into native space and applied to the participant’s pre-TMS scan. In each participant, the resultant connectivity maps yielded a correlation cluster in the left posterior inferior parietal lobule (IPL, Supplemental Figure 2). A target was placed in the averaged center of the left posterior IPL correlation cluster (formed from the overlay of the left posterior IPL clusters derived from connectivity maps) on the cortical surface using Brainsight neuronavigation software (Rogue Research, Inc.).

**Supplemental Tables**

Supplemental Table 1

| **Cue Reactive Network** | | **term** | **estimate** | **std.error** | **statistic** | **df** | **p-value** |  |
| --- | --- | --- | --- | --- | --- | --- | --- | --- |
| Main Model | |  |  |  |  |  |  |  |
|  |  | (Intercept) | 0.424 | 0.106 | 4.021 | 41.916 | 0.000 |  |
|  |  | diagnosisSchizophrenia | -0.009 | 0.072 | -0.127 | 56.806 | 0.899 |  |
|  |  | cueSmoke | 0.196 | 0.040 | 4.894 | 40.000 | 0.000 |  |
|  |  | demog_age | -0.002 | 0.003 | -0.686 | 39.000 | 0.497 |  |
|  |  | diagnosisSchizophrenia:cueSmoke | -0.009 | 0.063 | -0.148 | 40.000 | 0.883 |  |
|  |  | sd__(Intercept) | 0.169 | NA | NA | NA | NA |  |
|  |  | sd__Observation | 0.142 | NA | NA | NA | NA |  |
| Pairwise Comparison | |  |  |  |  |  |  |  |
|  | Control - Schizophrenia | Neutral-Baseline | 0.009 | 0.072 | 0.127 | 56.806 | 0.899 |  |
|  | Control - Schizophrenia | Nicotine-Baseline | 0.018 | 0.072 | 0.257 | 56.806 | 0.798 |  |
| **mPFC** |  | **term** | **estimate** | **std.error** | **statistic** | **df** | **p-value** | **FDRp** |
| Main Model | |  |  |  |  |  |  |  |
|  |  | (Intercept) | -0.015 | 0.137 | -0.111 | 43.082 | 0.912 | 0.912 |
|  |  | diagnosisSchizophrenia | -0.160 | 0.095 | -1.675 | 62.764 | 0.099 | 0.396 |
|  |  | cueSmoke | 0.171 | 0.061 | 2.811 | 40.000 | 0.008 | 0.008 |
|  |  | demog_age | -0.005 | 0.003 | -1.631 | 39.000 | 0.111 | 0.444 |
|  |  | diagnosisSchizophrenia:cueSmoke | 0.034 | 0.096 | 0.359 | 40.000 | 0.721 | 0.797 |
|  |  | sd__(Intercept) | 0.202 | NA | NA | NA | NA |  |
|  |  | sd__Observation | 0.216 | NA | NA | NA | NA |  |
| Pairwise Comparison | |  |  |  |  |  |  |  |
|  | Control - Schizophrenia | Neutral-Baseline | 0.160 | 0.095 | 1.675 | 62.764 | 0.099 | 0.396 |
|  | Control - Schizophrenia | Nicotine-Baseline | 0.125 | 0.095 | 1.314 | 62.764 | 0.194 | 0.598 |
| **PCC** |  | **term** | **estimate** | **std.error** | **statistic** | **df** | **p-value** | **FDRp** |
| Main Model | |  |  |  |  |  |  |  |
|  |  | (Intercept) | -0.159 | 0.143 | -1.107 | 44.284 | 0.274 | 0.365 |
|  |  | diagnosisSchizophrenia | -0.015 | 0.103 | -0.149 | 67.891 | 0.882 | 0.882 |
|  |  | cueSmoke | 0.259 | 0.072 | 3.589 | 40.000 | 0.001 | 0.001 |
|  |  | demog_age | -0.002 | 0.003 | -0.478 | 39.000 | 0.636 | 0.636 |
|  |  | diagnosisSchizophrenia:cueSmoke | -0.086 | 0.114 | -0.756 | 40.000 | 0.454 | 0.797 |
|  |  | sd__(Intercept) | 0.191 | NA | NA | NA | NA |  |
|  |  | sd__Observation | 0.255 | NA | NA | NA | NA |  |
| Pairwise Comparison | |  |  |  |  |  |  |  |
|  | Control - Schizophrenia | Neutral-Baseline | 0.015 | 0.103 | 0.149 | 67.891 | 0.882 | 0.882 |
|  | Control - Schizophrenia | Nicotine-Baseline | 0.101 | 0.103 | 0.984 | 67.891 | 0.329 | 0.598 |
| **L Lateral Occipital** | | **term** | **estimate** | **std.error** | **statistic** | **df** | **p-value** | **FDRp** |
| Main Model | |  |  |  |  |  |  |  |
|  |  | (Intercept) | 0.786 | 0.155 | 5.080 | 40.079 | 0.000 | 0 |
|  |  | diagnosisSchizophrenia | 0.047 | 0.099 | 0.474 | 45.890 | 0.638 | 0.882 |
|  |  | cueSmoke | 0.146 | 0.036 | 4.029 | 40.000 | 0.000 | 0 |
|  |  | demog_age | 0.002 | 0.004 | 0.512 | 39.000 | 0.612 | 0.636 |
|  |  | diagnosisSchizophrenia:cueSmoke | 0.029 | 0.057 | 0.506 | 40.000 | 0.616 | 0.797 |
|  |  | sd__(Intercept) | 0.277 | NA | NA | NA | NA |  |
| Pairwise Comparison | | sd__Observation | 0.128 | NA | NA | NA | NA |  |
|  | Control - Schizophrenia | Neutral-Baseline | -0.047 | 0.099 | -0.474 | 45.890 | 0.638 | 0.882 |
|  | Control - Schizophrenia | Nicotine-Baseline | -0.076 | 0.099 | -0.763 | 45.890 | 0.449 | 0.598 |
| **R Lateral Occipital** | | **term** | **estimate** | **std.error** | **statistic** | **df** | **p-value** | **FDRp** |
| Main Model | |  |  |  |  |  |  |  |
|  |  | (Intercept) | 1.155 | 0.203 | 5.687 | 40.046 | 0.000 | 0 |
|  |  | diagnosisSchizophrenia | 0.021 | 0.130 | 0.161 | 45.682 | 0.873 | 0.882 |
|  |  | cueSmoke | 0.190 | 0.047 | 4.076 | 40.000 | 0.000 | 0 |
|  |  | demog_age | -0.003 | 0.005 | -0.663 | 39.000 | 0.511 | 0.636 |
|  |  | diagnosisSchizophrenia:cueSmoke | 0.019 | 0.073 | 0.259 | 40.000 | 0.797 | 0.797 |
|  |  | sd__(Intercept) | 0.365 | NA | NA | NA | NA |  |
|  |  | sd__Observation | 0.165 | NA | NA | NA | NA |  |
| Pairwise Comparison | |  |  |  |  |  |  |  |
|  | Control - Schizophrenia | Neutral-Baseline | -0.021 | 0.130 | -0.161 | 45.682 | 0.873 | 0.882 |
|  | Control - Schizophrenia | Nicotine-Baseline | -0.040 | 0.130 | -0.307 | 45.682 | 0.760 | 0.760 |

Supplemental Table 2

| Cue Reactive Network  Pre-Treatment Nicotine>Neutral | | Estimate | SE | Statistic | P-value |
| --- | --- | --- | --- | --- | --- |
|  | Age | -0.004 | 0.002 | -1.940 | 0.059 |
|  | CPZ equivalents | 0.000 | 0.000 | -1.912 | 0.076 |
|  | BPRS total | 0.001 | 0.003 | 0.366 | 0.715 |
|  | BPRS negative | 0.005 | 0.012 | 0.482 | 0.632 |
| mPFC  Pre-Treatment Nicotine>Neutral | | Estimate | SE | Statistic | P-value |
|  | Age | -0.002 | 0.003 | -0.606 | 0.547 |
|  | CPZ equivalents | 0.000 | 0.000 | -0.931 | 0.367 |
|  | BPRS total | 0.006 | 0.005 | 1.361 | 0.181 |
|  | BPRS negative | 0.028 | 0.018 | 1.540 | 0.131 |
| PCC  Pre-Treatment Nicotine>Neutral | | Estimate | SE | Statistic | P-value |
|  | Age | -0.005 | 0.004 | -1.361 | 0.181 |
|  | CPZ equivalents | 0.000 | 0.000 | -0.931 | 0.367 |
|  | BPRS total | 0.004 | 0.006 | 0.730 | 0.469 |
|  | BPRS negative | 0.018 | 0.022 | 0.835 | 0.408 |
| L Lateral Occipital  Pre-Treatment Nicotine>Neutral | | Estimate | SE | Statistic | P-value |
|  | Age | -0.004 | 0.002 | -2.006 | 0.051 |
|  | CPZ equivalents | 0.000 | 0.000 | -1.522 | 0.150 |
|  | BPRS total | -0.001 | 0.002 | -0.415 | 0.679 |
|  | BPRS negative | -0.010 | 0.010 | -0.935 | 0.355 |
| R Lateral Occipital  Pre-Treatment Nicotine>Neutral | | Estimate | SE | Statistic | P-value |
|  | Age | -0.005 | 0.002 | -1.910 | 0.063 |
|  | CPZ equivalents | 0.000 | 0.000 | -1.280 | 0.221 |
|  | BPRS total | -0.003 | 0.003 | -0.879 | 0.384 |
|  | BPRS negative | -0.006 | 0.014 | -0.465 | 0.643 |

Supplemental Table 3

| **Cue Reactive Network Nicotine-Neutral contrast change pre/post TMS** | **term** | **estimate** | **std.error** | **statistic** | **df** | **p-value** |  |
| --- | --- | --- | --- | --- | --- | --- | --- |
| Main Model |  |  |  |  |  |  |  |
|  | (Intercept) | 0.150 | 0.110 | 1.363 | 56.000 | 0.178 |  |
|  | diagnosisSchizophrenia | 0.068 | 0.082 | 0.822 | 56.000 | 0.415 |  |
|  | tms_typeDMN | -0.039 | 0.072 | -0.547 | 56.000 | 0.587 |  |
|  | pre_reactivity | -0.924 | 0.149 | -6.207 | 56.000 | 0.000 |  |
|  | demog_age | -0.001 | 0.002 | -0.279 | 56.000 | 0.781 |  |
|  | diagnosisSchizophrenia:tms_typeDMN | -0.196 | 0.117 | -1.678 | 56.000 | 0.099 |  |
|  | sd__(Intercept) | 0.000 | NA | NA | NA | NA |  |
|  | sd__Observation | 0.222 | NA | NA | NA | NA |  |
| Estimated Marginal Means |  |  |  |  |  |  |  |
|  | Control_DLPFC | -0.013 | 0.051 | -0.249 | 55.947 | 0.805 |  |
|  | Schizophrenia_DLPFC | 0.055 | 0.065 | 0.846 | 55.952 | 0.401 |  |
|  | Control_DMN | -0.052 | 0.052 | -1.000 | 55.984 | 0.322 |  |
|  | Schizophrenia_DMN | -0.180 | 0.067 | -2.698 | 55.998 | 0.009 |  |
| **mPFC Nicotine-Neutral contrast change pre/post TMS** | **term** | **estimate** | **std.error** | **statistic** | **df** | **p-value** | **FDRp** |
| Main Model |  |  |  |  |  |  |  |
|  | (Intercept) | 0.105 | 0.125 | 0.837 | 56.000 | 0.406 | 0.416 |
|  | diagnosisSchizophrenia | -0.034 | 0.099 | -0.349 | 56.000 | 0.729 | 0.729 |
|  | tms_typeDMN | -0.093 | 0.086 | -1.079 | 56.000 | 0.285 | 0.834 |
|  | pre_reactivity | -0.914 | 0.128 | -7.115 | 56.000 | 0.000 | 0 |
|  | demog_age | 0.003 | 0.003 | 1.192 | 56.000 | 0.238 | 0.599 |
|  | diagnosisSchizophrenia:tms_typeDMN | -0.101 | 0.141 | -0.717 | 56.000 | 0.477 | 0.477 |
|  | sd__(Intercept) | 0.000 | NA | NA | NA | NA |  |
|  | sd__Observation | 0.265 | NA | NA | NA | NA |  |
| Estimated Marginal Means |  |  |  |  |  |  |  |
|  | Control_DLPFC | 0.084 | 0.061 | 1.364 | 55.987 | 0.178 | 0.356 |
|  | Schizophrenia_DLPFC | 0.049 | 0.078 | 0.632 | 55.997 | 0.530 | 0.865 |
|  | Control_DMN | -0.010 | 0.062 | -0.153 | 55.998 | 0.879 | 0.879 |
|  | Schizophrenia_DMN | -0.145 | 0.080 | -1.803 | 55.982 | 0.077 | 0.077 |
| **PCC  Nicotine-Neutral contrast change pre/post TMS** | **term** | **estimate** | **std.error** | **statistic** | **df** | **p-value** | **FDRp** |
| Main Model |  |  |  |  |  |  |  |
|  | (Intercept) | 0.264 | 0.160 | 1.651 | 40.572 | 0.106 | 0.416 |
|  | diagnosisSchizophrenia | 0.107 | 0.118 | 0.900 | 55.549 | 0.372 | 0.729 |
|  | tms_typeDMN | -0.021 | 0.101 | -0.212 | 26.308 | 0.834 | 0.834 |
|  | pre_reactivity | -1.048 | 0.131 | -7.977 | 55.713 | 0.000 | 0 |
|  | demog_age | -0.002 | 0.004 | -0.530 | 33.308 | 0.599 | 0.599 |
|  | diagnosisSchizophrenia:tms_typeDMN | -0.367 | 0.164 | -2.243 | 30.185 | 0.032 | 0.128 |
|  | sd__(Intercept) | 0.064 | NA | NA | NA | NA |  |
|  | sd__Observation | 0.308 | NA | NA | NA | NA |  |
| Estimated Marginal Means |  |  |  |  |  |  |  |
|  | Control_DLPFC | 0.036 | 0.074 | 0.485 | 55.770 | 0.630 | 0.630 |
|  | Schizophrenia_DLPFC | 0.142 | 0.092 | 1.539 | 55.855 | 0.129 | 0.516 |
|  | Control_DMN | 0.014 | 0.074 | 0.194 | 55.962 | 0.847 | 0.879 |
|  | Schizophrenia_DMN | -0.246 | 0.095 | -2.589 | 55.994 | 0.012 | 0.036 |
| **LLO Nicotine-Neutral contrast change pre/post TMS** | **term** | **estimate** | **std.error** | **statistic** | **df** | **p-value** | **FDRp** |
| Main Model |  |  |  |  |  |  |  |
|  | (Intercept) | 0.087 | 0.106 | 0.819 | 56.000 | 0.416 | 0.416 |
|  | diagnosisSchizophrenia | 0.054 | 0.081 | 0.666 | 56.000 | 0.508 | 0.729 |
|  | tms_typeDMN | -0.034 | 0.072 | -0.468 | 56.000 | 0.642 | 0.834 |
|  | pre_reactivity | -0.811 | 0.162 | -5.017 | 56.000 | 0.000 | 0 |
|  | demog_age | -0.002 | 0.002 | -0.649 | 56.000 | 0.519 | 0.599 |
|  | diagnosisSchizophrenia:tms_typeDMN | -0.098 | 0.116 | -0.841 | 56.000 | 0.404 | 0.477 |
|  | sd__(Intercept) | 0.000 | NA | NA | NA | NA |  |
|  | sd__Observation | 0.220 | NA | NA | NA | NA |  |
| Estimated Marginal Means |  |  |  |  |  |  |  |
|  | Control_DLPFC | -0.084 | 0.051 | -1.645 | 55.990 | 0.106 | 0.356 |
|  | Schizophrenia_DLPFC | -0.030 | 0.065 | -0.457 | 55.968 | 0.649 | 0.865 |
|  | Control_DMN | -0.118 | 0.052 | -2.250 | 55.966 | 0.028 | 0.112 |
|  | Schizophrenia_DMN | -0.161 | 0.066 | -2.433 | 56.000 | 0.018 | 0.036 |
| **RLO Nicotine-Neutral contrast change pre/post TMS** | **term** | **estimate** | **std.error** | **statistic** | **df** | **p-value** | **FDRp** |
| Main Model |  |  |  |  |  |  |  |
|  | (Intercept) | 0.160 | 0.141 | 1.135 | 34.890 | 0.264 | 0.416 |
|  | diagnosisSchizophrenia | 0.060 | 0.104 | 0.581 | 55.157 | 0.563 | 0.729 |
|  | tms_typeDMN | -0.039 | 0.086 | -0.453 | 23.365 | 0.655 | 0.834 |
|  | pre_reactivity | -0.826 | 0.176 | -4.703 | 54.012 | 0.000 | 0 |
|  | demog_age | -0.002 | 0.003 | -0.624 | 30.199 | 0.537 | 0.599 |
|  | diagnosisSchizophrenia:tms_typeDMN | -0.147 | 0.139 | -1.057 | 27.018 | 0.300 | 0.477 |
|  | sd__(Intercept) | 0.109 | NA | NA | NA | NA |  |
|  | sd__Observation | 0.259 | NA | NA | NA | NA |  |
| Estimated Marginal Means |  |  |  |  |  |  |  |
|  | Control_DLPFC | -0.052 | 0.065 | -0.800 | 55.455 | 0.427 | 0.569 |
|  | Schizophrenia_DLPFC | 0.009 | 0.082 | 0.104 | 55.650 | 0.917 | 0.917 |
|  | Control_DMN | -0.091 | 0.067 | -1.366 | 55.463 | 0.178 | 0.356 |
|  | Schizophrenia_DMN | -0.178 | 0.084 | -2.113 | 55.878 | 0.039 | 0.052 |

Supplemental Table 4

|  | TMS-Order effect | SE | Statistic | P-value | FDRp |
| --- | --- | --- | --- | --- | --- |
| Cue Reactive Network | 0.004 | 0.061 | 0.068 | 0.945 |  |
| mPFC | 0.010 | 0.073 | 0.142 | 0.886 | 0.886 |
| PCC | 0.066 | 0.088 | 0.758 | 0.454 | 0.886 |
| L Lateral Occipital | -0.025 | 0.061 | -0.423 | 0.673 | 0.886 |
| R Lateral Occipital | -0.053 | 0.081 | -0.653 | 0.519 | 0.886 |

Supplemental Table 5

|  |  | Change in Nicotine>Neutral Cue-Reactivity | SE | Statistic | P-value | FDRp |
| --- | --- | --- | --- | --- | --- | --- |
| During Washout Period in SZ | |  |  |  |  |  |
|  | Cue Reactive Network | -0.034 | 0.075 | -0.450 | 0.653 |  |
|  | mPFC | 0.030 | 0.106 | 0.283 | 0.777 | 0.897 |
|  | PCC | -0.016 | 0.126 | -0.129 | 0.897 | 0.897 |
|  | L Lateral Occipital | -0.080 | 0.064 | -1.235 | 0.220 | 0.732 |
|  | R Lateral Occipital | -0.079 | 0.087 | -0.909 | 0.366 | 0.732 |
| During Washout Period in NC | |  |  |  |  |  |
|  | Cue Reactive Network |  |  |  |  |  |
|  | mPFC | 0.124 | 0.079 | 1.560 | 0.123 | 0.246 |
|  | PCC | 0.058 | 0.094 | 0.617 | 0.539 | 0.539 |
|  | L Lateral Occipital | -0.115 | 0.047 | -2.419 | 0.018 | 0.072 |
|  | R Lateral Occipital | -0.062 | 0.065 | -0.967 | 0.336 | 0.448 |
| Between Two TMS Start Dates in SZ | |  |  |  |  |  |
|  | Cue Reactive Network | 0.124 | 0.079 | 1.560 | 0.121 |  |
|  | mPFC | 0.167 | 0.101 | 1.655 | 0.101 | 0.404 |
|  | PCC | 0.047 | 0.120 | 0.389 | 0.697 | 0.694 |
|  | L Lateral Occipital | 0.066 | 0.061 | 1.084 | 0.281 | 0.498 |
|  | R Lateral Occipital | 0.074 | 0.083 | 0.893 | 0.374 | 0.498 |
| Between Two TMS Start Dates in NC | |  |  |  |  |  |
|  | Cue Reactive Network | 0.059 | 0.054 | 1.078 | 0.284 |  |
|  | mPFC | 0.055 | 0.077 | 0.722 | 0.471 | 0.856 |
|  | PCC | 0.029 | 0.062 | 0.466 | 0.642 | 0.856 |
|  | L Lateral Occipital | 0.002 | 0.046 | 0.062 | 0.950 | 0.95 |
|  | R Lateral Occipital | 0.029 | 0.062 | 0.466 | 0.642 | 0.856 |

Supplemental Table 6

| **Cue Reactive Network [Nicotine-Neutral] Change** | **term** | **estimate** | **std.error** | **statistic** | **df** | **p-value** |  |
| --- | --- | --- | --- | --- | --- | --- | --- |
|  | (Intercept) | 0.241041945 | 0.108089676 | 2.230018201 | 55 | 0.029846999 |  |
|  | BaselineCraving | -0.033756869 | 0.011172343 | -3.021467157 | 55 | 0.003813043 |  |
|  | pre_reactivity | -0.837164208 | 0.152073888 | -5.504983269 | 55 | 1.00E-06 |  |
|  | demog_age | -0.000340969 | 0.002372411 | -0.143722605 | 55 | 0.886244826 |  |
|  | sd__(Intercept) | 0 | NA | NA | NA | NA |  |
|  | sd__Observation | 0.21885072 | NA | NA | NA | NA |  |
| **mPFC [Nicotine-Neutral] Change** | **term** | **estimate** | **std.error** | **statistic** | **df** | **p-value** | **FDRp** |
|  | (Intercept) | 0.165325932 | 0.121378342 | 1.362071103 | 55 | 0.178728118 | 0.178 |
|  | BaselineCraving | -0.039964955 | 0.013021679 | -3.069109118 | 55 | 0.003330621 | 0.006 |
|  | pre_reactivity | -0.77344228 | 0.130124557 | -5.943861015 | 55 | 1.98E-07 | 0 |
|  | demog_age | 0.003882534 | 0.002722241 | 1.426227351 | 55 | 0.159454501 | 0.593 |
|  | sd__(Intercept) | 0 | NA | NA | NA | NA |  |
|  | sd__Observation | 0.253758336 | NA | NA | NA | NA |  |
| **PCC [Nicotine-Neutral] Change** | **term** | **estimate** | **std.error** | **statistic** | **df** | **p-value** | **FDRp** |
|  | (Intercept) | 0.302608834 | 0.162496228 | 1.862251433 | 55 | 0.067912942 | 0.090 |
|  | BaselineCraving | -0.034376206 | 0.016216859 | -2.11978207 | 55 | 0.038551748 | 0.038 |
|  | pre_reactivity | -0.974236393 | 0.140056353 | -6.956031406 | 55 | 4.45E-09 | 0 |
|  | demog_age | -7.55E-08 | 0.003558475 | -2.12E-05 | 55 | 0.999983148 | 0.99 |
|  | sd__(Intercept) | 0 | NA | NA | NA | NA |  |
|  | sd__Observation | 0.321270325 | NA | NA | NA | NA |  |
| **LLO [Nicotine-Neutral] Change** | **term** | **estimate** | **std.error** | **statistic** | **df** | **p-value** | **FDRp** |
|  | (Intercept) | 0.238389733 | 0.100254912 | 2.377835957 | 55 | 0.020917267 | 0.083 |
|  | BaselineCraving | -0.034201847 | 0.010522354 | -3.250398929 | 55 | 0.001969338 | 0.006 |
|  | pre_reactivity | -0.762992433 | 0.150531546 | -5.068654715 | 55 | 4.85E-06 | 0 |
|  | demog_age | -0.002334675 | 0.002217368 | -1.052903441 | 55 | 0.296989123 | 0.593 |
|  | sd__(Intercept) | 0 | NA | NA | NA | NA |  |
|  | sd__Observation | 0.206295521 | NA | NA | NA | NA |  |
| **RLO [Nicotine-Neutral] Change** | **term** | **estimate** | **std.error** | **statistic** | **df** | **p-value** | **FDRp** |
|  | (Intercept) | 0.266411023 | 0.137087667 | 1.943362438 | 40.37366429 | 0.058966511 | 0.090 |
|  | BaselineCraving | -0.032990094 | 0.014666411 | -2.249363792 | 49.94122202 | 0.028929371 | 0.038 |
|  | pre_reactivity | -0.756371069 | 0.175832879 | -4.301647529 | 53.59080435 | 7.22E-05 | 0 |
|  | demog_age | -0.002162295 | 0.003102849 | -0.696874255 | 32.5921689 | 0.490822487 | 0.654 |
|  | sd__(Intercept) | 0.108230218 | NA | NA | NA | NA |  |
|  | sd__Observation | 0.253950548 | NA | NA | NA | NA |  |

**Supplemental Figures**

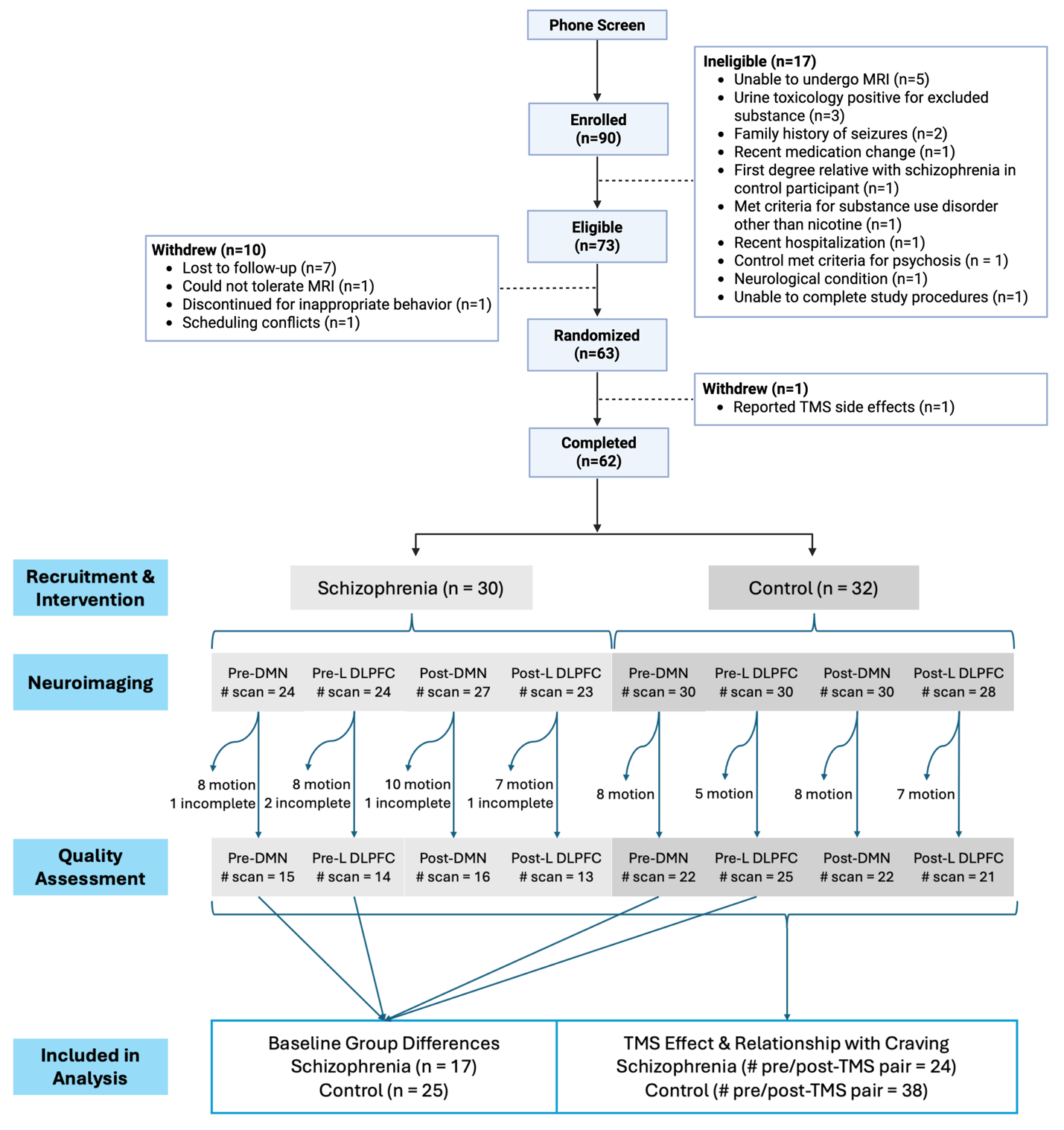

**Supplemental Figure 1. Consort Diagram**. Sixty-two participants (30 schizophrenia, 32 non-psychosis control) completed the study and provided data for analysis. 216 task functional magnetic resonance imaging (fMRI) scans (98 schizophrenia, 118 non-psychosis control) were acquired. 148 task fMRI scans passed quality assessment (58 schizophrenia, 90 non-psychosis control) were included in data analysis. Created with BioRender.com.

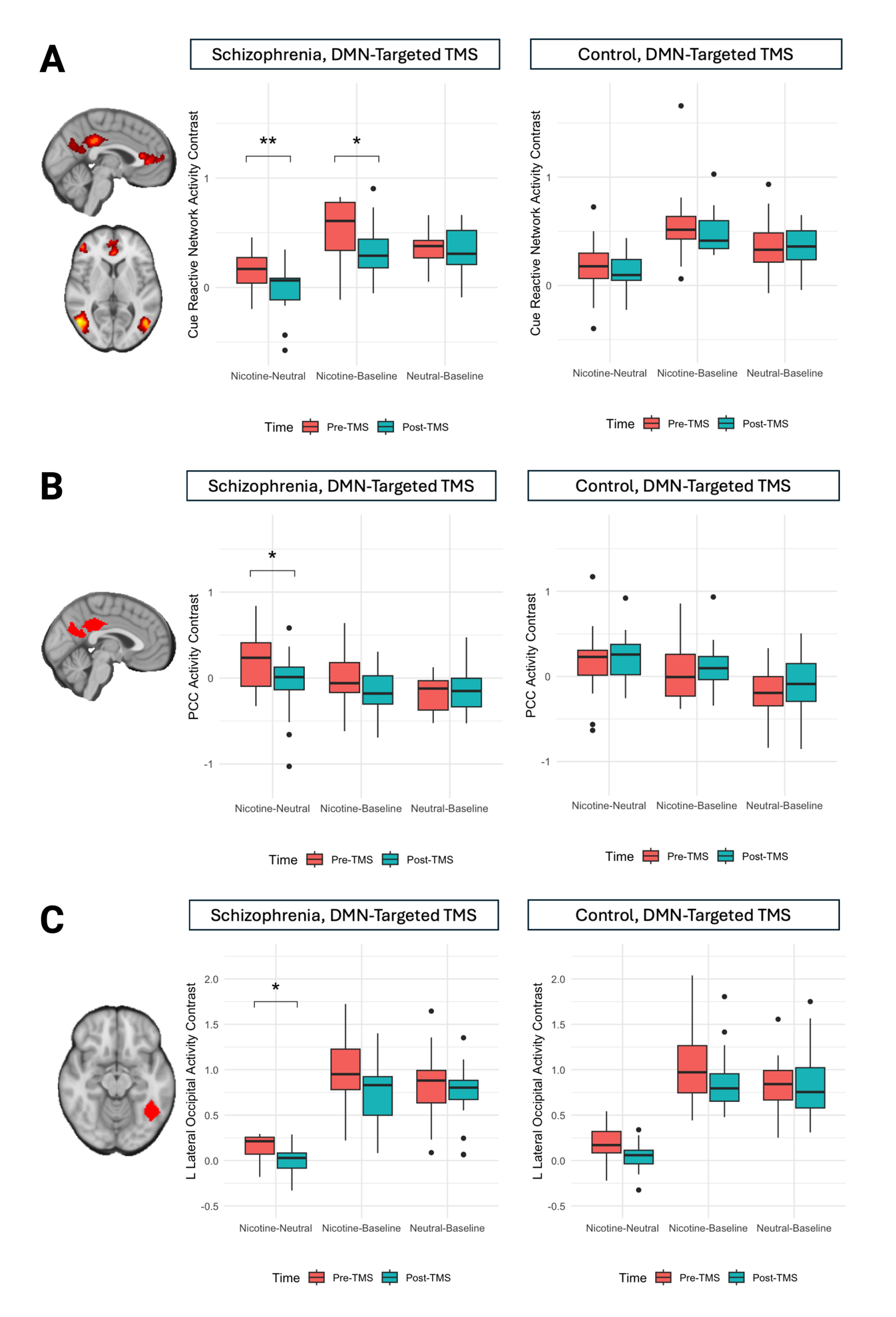

**Supplemental Figure 2. DMN-Targeted TMS Decreased Cue Reactivity in SZ Across the Cue Reactive Network.** We modeled TMS-induced cue-reactivity change using diagnosis-by-TMS-type interaction, controlling for age. Estimated marginal means indicated that only the SZ/DMN-targeted-TMS group exhibited significant changes in cue-reactivity. In SZ, DMN-targeted TMS significantly decreased Nicotine>Neutral contrast in CRN (estimate=-0.180, p=0.009; Figure 3A), PCC (estimate=-0.26, p=0.012, FDRp=0.013; Figure 3B), and LLO (estimate=-0.161, p=0.018, FDRp=0.025; Figure 3C). Decrease in Nicotine>Neutral contrast in CRN was driven by decreased reactivity to Nicotine cues (Nicotine>Baseline: estimate=-0.135, p=0.035; Figure 3A). DMN-targeted TMS did not change reactivity to Neutral cues. Created with BioRender.com.

1.	First M, Williams J, Karg R, Spitzer R. *Structured Clinical Interview for DSM-5—Research Version (SCID-5 for DSM-5, Research Version; SCID-5-RV).* American Psychiatric Association; 2015.

2.	Fox MD, Buckner RL, White MP, Greicius MD, Pascual-Leone A. Efficacy of Transcranial Magnetic Stimulation Targets for Depression Is Related to Intrinsic Functional Connectivity with the Subgenual Cingulate. *Biol Psychiatry*. 2012;72(7):595-603. doi:10.1016/j.biopsych.2012.04.028

3.	Huang YZ, Edwards MJ, Rounis E, Bhatia KP, Rothwell JC. Theta Burst Stimulation of the Human Motor Cortex. *Neuron*. 2005;45(2):201-206. doi:10.1016/j.neuron.2004.12.033

4.	Eldaief MC, Halko MA, Buckner RL, Pascual-Leone A. Transcranial magnetic stimulation modulates the brain’s intrinsic activity in a frequency-dependent manner. *Proc Natl Acad Sci*. 2011;108(52):21229-21234. doi:10.1073/pnas.1113103109

5.	Thomas Yeo BT, Krienen FM, Sepulcre J, et al. The organization of the human cerebral cortex estimated by intrinsic functional connectivity. *J Neurophysiol*. 2011;106(3):1125-1165. doi:10.1152/jn.00338.2011
